# A pragmatic pipeline for drug resistance identification in *Mycobacterium tuberculosis* using whole genome sequencing

**DOI:** 10.1101/2024.04.15.24305720

**Authors:** Linzy Elton, Alp Aydin, Neil Stoker, Sylvia Rofael, Letícia Muraro Wildner, Jabar Babatunde Pacome Agbo Achimi Abdul, John Tembo, Muzamil Abdel Hamid, Mfoutou Mapanguy Claujens Chastel, Julio Ortiz Canseco, Ronan Doyle, Giovanni Satta, Justin O’Grady, Adam Witney, Francine Ntoumi, Alimuddin Zumla, Timothy D McHugh

**Affiliations:** Centre for Clinical Microbiology, University College London, UK; Oxford Nanopore Technologies plc. Quadram Institute, Rosalind Franklin Road, Norwich Research Park, Norwich, UK; Faculty of Pharmacy, Alexandria University, Egypt; Centre de Recherches Médicales de Lambaréné, Gabon; HerpeZ, University Teaching Hospital, Lusaka, Zambia; Institute for Endemic Diseases, University of Khartoum, Khartoum, Sudan; Fondation Congolaise pour la Recherche Médicale, Republic of Congo; Université Marien Ngouabi, Brazzaville Republic of Congo; Francis Crick Institute, London, UK; London School of Hygiene and Tropical Medicine, University of London, London, UK; University College London Hospitals NHS Foundation Trust, London, UK; St George’s, University of London, London, UK; National Institute for Health Research Biomedical Research Centre, University College London Hospitals NHS Foundation Trust, London, UK

**Keywords:** tuberculosis, drug resistance, whole genome sequencing, Oxford Nanopore, MinION, diagnosis

## Abstract

**Background:** Delays in accurate diagnosis of drug resistant tuberculosis (DR-TB) can hinder treatment. Whole genome sequencing (WGS) provides more information than standard molecular and phenotypic testing, but commonly used platforms are expensive to implement, and data interpretation requires significant expertise.

**Aims:** We aimed to optimise a TB WGS diagnostic pipeline balancing user-friendliness, cost- effectiveness and time to results, whilst ensuring accuracy.

**Materials and methods:** Growth conditions, DNA extraction protocols and Oxford Nanopore Technologies (ONT) library preparation kits were compared. Software for basecalling and analysis were evaluated to find the most accurate resistance SNP and lineage predictor.

**Results:** Optimally, a spin-column CTAB DNA extraction method was combined with the RBK110.96 library preparation kit, high accuracy basecalling and data analysis using TB-Profiler. Compared with Illumina, the pipeline was concordant for 16/17 (94%) isolates (lineage) and for 17/17 (100%) isolates (resistance SNPs). Our pipeline was 71% (12/17) concordant with phenotypic drug susceptibility test (DST) results. Time-to-diagnosis was around four weeks.

**Conclusions:** This optimised TB sequencing pipeline requires less time expertise to run and analyse than Illumina, takes less time than phenotypic DSTs and the results are comparable with Illumina. The cost per sample is comparable with other methods. These features make it an important tool for incorporating into routine DR-TB diagnostic pipelines and larger scale drug resistance surveillance in all settings.

## Introduction

Treatment for drug-resistant tuberculosis (DR-TB) is intensive and can take up to 20 months to complete, compliance can be low due to drug toxicity, and there is increased risk of mortality (1). Misdiagnosis and delays to correct diagnosis, both between drug sensitive (DS-TB), DR-TB and multi- drug resistant (MDR-TB) strains, and the differentiation between *M. tuberculosis* and non- tuberculosis mycobacteria (NTMs), can complicate and delay appropriate treatment. Delayed treatment is associated with high early mortality and the increase in drug resistance prevalence (2). Whilst the prevalence of DR-TB has remained stable over the last decade, the top 30 countries with the highest TB prevalence identified by WHO are low- and middle-income countries (LMICs) (3). DR- TB has been most commonly reported from Southern America and Central Asia, although data is not available from many African countries (4,5). Phenotypic drug sensitivity testing (DST) tends to be restricted to reference facilities or specialist centres, but the roll out of molecular assays such as the GeneXpert MTB/RIF Ultra and MTB/XDR (Cepheid) or the line probe assay GenoType® MTBDR (Bruker Corporation) has allowed more localised testing. However, these are incomprehensive targeted methods which are limited to detecting specific resistance-encoding mutations to certain drugs (6,7).

Whole genome sequencing (WGS) has emerged as a diagnostic tool for DR-TB cases (8–10). WGS is a comprehensive molecular diagnostic test that can identify mutations for all drugs in one assay including novel drug resistance mutations and novel drug targets. It can predict resistance through gene inactivation caused by multiple mutations, frameshift mutations or large structural variations in the genome [9]. WGS can also identify TB lineages and mixed strain infections (11). In outbreak situations, WGS is a powerful tool to identify transmission chains and clusters (12). In cases of negative treatment outcomes, WGS can differentiate between cases of relapse, due to treatment failure, and reinfection, which might be due to failure to develop protective immunity, through calculating the genetic distance between pre- and post-treatment *M. tuberculosis* genomes (13). Despite the increasing availability of sequencing platforms in LMICs, the lack of bioinformaticians, data analysis training opportunities, and high-performance computing facilities have been a bottleneck to the roll out of WGS globally (14).

Despite the potential of WGS, *M. tuberculosis* is a challenging organism to work with. DNA extraction is complicated by the complex cell wall structure (15). Many laboratories utilise the cethyl trimethyl ammonium bromide (CTAB) method (16), which generally produces a higher yield of genomic DNA of good integrity and purity for sequencing applications compared to commercial kits, and the reagents for which are mostly available and commonly used in TB laboratories in LMICs. DR-TB isolates tend to be slower to grow than DS-TB ones and generally produce less biomass, which can make extracting sufficient amounts of genomic DNA more difficult (17).

The mycobacterial genome has a high GC content (>60%) and multiple repeat regions. These structural features create challenges when considering sequencing library preparation and data analysis (18,19). On the other hand, an advantage in the analysis of the DR genotype for *M. tuberculosis* is that mycobacterial resistance is mainly dependent on the evolution of mutations such as single nucleotide polymorphisms (SNPs) and insertions or deletions (INDELs) within chromosomal genes rather than horizontal gene transmission (20).

The employment of short read, high throughput sequencing platforms, such as the Illumina MiSeq, is established for DR-TB genotyping (21–23), but these platforms have high resource requirements and highly skilled operators, and in consequence such large scale sequencing is not always available, especially in resource-constrained settings. In contrast, long read sequencing offered by ONT using the MinION; a small, portable device, is accessible to resource-constrained environments because of lower set-up costs and less complex training needs. Long read sequencing can additionally be advantageous for sequencing complex genomes which are GC rich or have many repeated regions.

There is little research from resource-constrained settings on the use of ONT platforms for TB diagnosis. Those publishing have often done so as individual research studies (24–29). Previous studies often made use of the ligation-based ONT library preparation (25,28). These require costly third-party reagents and high DNA input (1 µg), which can be hard to routinely obtain. If optimised appropriately, use of the rapid barcoding kits, employing multiplexing, should improve cost effectiveness (24).

Whilst phenotypic DSTs are the benchmark and used as a standard as there are international standards, diagnosis utilising WGS may help to provide more accurate results and therefore a faster correct drug regimen. Genomics is especially important in countries with no DST capabilities, or for new drugs like bedaquiline, where there is no other diagnostic. Currently, there does not appear to be a TB sequencing protocol that could be integrated into diagnostic pipelines in resource constrained settings to aid DR-TB diagnosis. We aimed to develop a pipeline, optimising methodology to create a tool that had an optimal combination of accuracy, speed, cost-effectiveness and user-friendliness.

## Materials and methods

The full manual for this pipeline, including protocols and tutorial videos can be found on the PANDORA-ID-NET Global Health Network hub (30).

### M. tuberculosis samples and ethics statement

17 *M. tuberculosis* isolates from an archive maintained by the Centre for Clinical Microbiology (CCM) at University College London (UCL), which have metadata and phenotypic drug sensitivity data (streptomycin, isoniazid, rifampicin, ethambutol plus pyrazinamide) were utilised. Eight DS-TB isolates, five isoniazid mono-resistant isolates and four MDR-TB isolates were selected. The study did not include live participants. The study used archived *M. tuberculosis* isolates and no patient data was included in the study. There is no means to associate the isolates with the original source and ethical approval was not required.

### Growth of M. tuberculosis

*M. tuberculosis* isolates were incubated in MGIT tubes (Becton Dickson), using the BD BACTEC™ MGIT™ automated mycobacterial detection system or on Middlebrook 7H11 slopes (EO laboratories) and incubated at 37°C in a static incubator. To identify the optimal growth time to achieve the greatest DNA yield, two representative MDR-TB and two DS-TB samples were selected from the collection. 500 µL of MGIT subcultures were seeded onto 7H11 slopes and into MGIT tubes and left to grow. Once the MGIT flagged positive, each isolate was further cultured for 2, 3 or 4 weeks, after which DNA was extracted.

### DNA extraction

Two different CTAB methods were compared. For each extraction method, 400 µL of liquid culture taken from a MGIT tube was used.

Method 1: The precipitation CTAB DNA extraction method, as described in Kent *et al* . (16) was optimised, with the addition of a 15-minute sonication step after heat killing and 3 µL yeast tRNA (Fisher Scientific) at the isopropanol precipitation step. DNA was eluted into 25 µL molecular grade water.

Method 2: A spin-column CTAB method, developed by ONT, was compared. Briefly, samples were heat killed, then glass beads and lysozyme (10mg/mL) added. The samples were incubated in a shaking incubator at 37°C for 15 min at 800 RPM. Proteinase K and CTAB buffer were added and then incubated at 56°C for 30 min at 1000 RPM. 5M NaCl was added and the debris pelleted by centrifugation, before a modified version of the Monarch Genomic Purification Kit (New England Biosciences) was followed. The full protocol for spin-column CTAB is in supplementary materials S1.

### DNA quantification

DNA quantified using spectrophotometry and the Qubit™ dsDNA BR Assay Kit (Thermo Fisher) and DNA size and integrity confirmed using electrophoresis with the Genomic DNA ScreenTape and reagents on the TapeStation 4150 (Agilent Technologies Inc.).

### DNA shearing

DNA was sheared mechanically using g-TUBES (Covaris), following manufacturer’s instructions (31). Samples with an average size of 10 kb, 20 kb and ∼50 kb were sequenced for 2 hours each and the output number of reads, N50 (the sequence length of the shortest contig at 50% coverage of the total genome assembly) and bases compared.

### Library preparation

DNA libraries were prepared using either the Rapid Barcoding Kit (SQK-RBK004) (ONT) with a DNA input of ∼400 ng, the newer Rapid Barcoding Kit 96 (SQK-RBK110.96) (ONT) with a DNA input of between 50-200 ng, or the PCR Barcoding kit (SQK-RPB004) (ONT) with DNA inputs of 1-5 ng, following the respective ONT kit protocols (32–34). DNA libraries were prepared from the same spin-column DNA extractions for Illumina sequencing using the DNA Prep, (M) Tagmentation (24 Samples, IPB) library preparation kit (Illumina) and the MiSeq Reagent Kit v3 (600-cycle) (Illumina) (35,36).

### Sequencing and basecalling

Twelve or 24 barcoded samples were run together on a flow cell version R9.4.1 (ONT) using a MinION device for 72 hours, using the default parameters on the MinKNOW software (v23.04.6). Basecalling was performed either by the MinKNOW software alongside sequencing or using the Guppy basecalling software (v6.5.7) (ONT) (37), using either the flip-flop Fast, high accuracy (HAC), or super-high accuracy (SUP) algorithm. Basecalling was undertaken on a 32 GB RAM CPU computer (for specifications, see Supplementary materials S2).

### Data analysis

Fastq files were quality checked (QC) using FastQC (v0.21.1) and MultiQC (v1.15) (38,39). Barcodes were demultiplexed and trimmed from the reads using Guppy. To view depth and coverage of the sequences, fastq files were aligned to the H37Rv reference genome (NCBI Reference Sequence NC_000962.3) using minimap2 (v2.26) (40), then sorted and indexed using SAMtools (v1.17) (41). Alignments were visualised using Artemis (v18.1.0) (42). Depth and coverage were calculated using SAMtools. Fastq files were input to the resistance gene SNP and lineage calling software configured for ONT data, Mykrobe (v0.10.0) (43) and TB-Profiler (v4.4.0) (44) graphical user interfaces (GUIs) to compare accuracy. ONT results were compared to Illumina data obtained from the same extraction. Drug resistance SNPs with a confidence grading of ≥3, as defined by the WHO were included in the comparison (45). Data were analysed using Prism (v10.0.0.153) (GraphPad). Sequence data were deposited under Project accession number PRJEB68143 on the European Nucleotide Archive and outlined in supplementary materials S3.

## Results

DNA yield and fragment size for cultures sampled over time are displayed in Figure 1. The mean DNA yield for the MDR-TB isolates was 688 ng (SD=413) (from MGIT liquid culture) and 966 ng (SD=841) (from 7H11 slopes), whilst for the DS-TB isolates it was 1414 ng (SD=1265) (MGIT) and 1712 ng (SD=1327) (7H11). The quantity of DNA obtained from 7H11 was significantly higher than from MGITs especially for the MDR-TB isolates (p=<0.0001, unpaired t-test). For MDR-TB isolates, both the MGIT and 7H11 yields did not change significantly over time. For DS-TB isolates, for both the MGIT and 7H11 cultures, the yields decreased over time.

**Figure 1.**
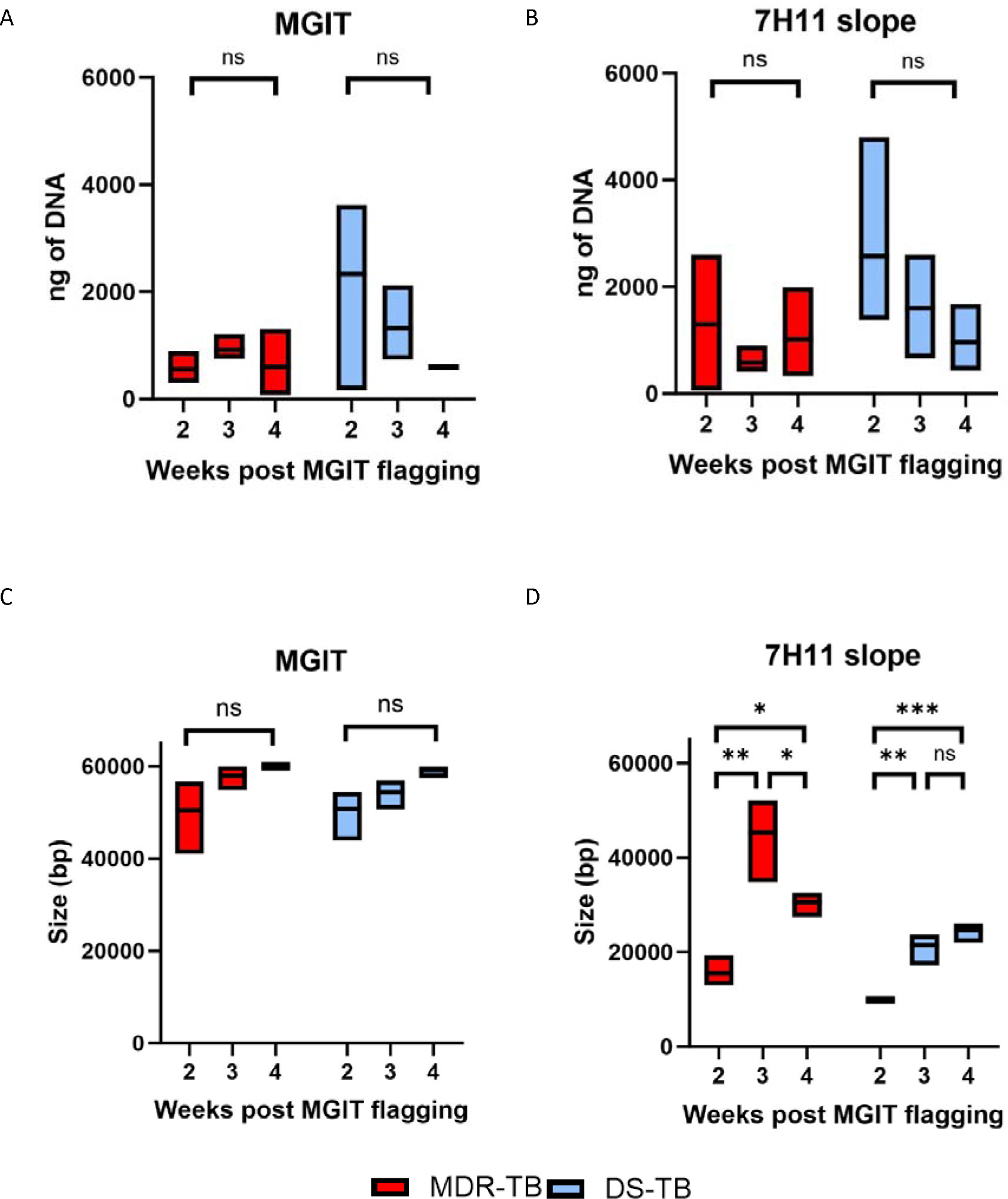
Genomic DNA yield and integrity obtained from MDR-TB and DS-TB isolate cultures. Overall, 7H11 slopes produced the greatest yield of DNA, whilst MGIT tubes produced the greatest DNA size. A) DNA yields extracted from MGIT tubes over time; there was no change in MDR-TB DNA yield, but the DS-TB isolate yield decreased. B) DNA amounts extracted from 7H11 slopes over time; there was no change in MDR-TB DNA yields, but DS-TB isolate yields decreased. C) Size of DNA extracted from MGIT tubes over time; both MDR-TB and DS-TB isolates increased over time. D) Size of DNA extracted from 7H11 slopes over time; the MDR-TB isolates peaked at week 3 and then decreased (p=0.0025), the DS-TB isolates peaked at weeks 3 and 4 (p=0.01). Significance was calculated using One-Way ANOVA and Tukey’s post ad hoc analysis.

The largest DNA fragments for both MDR-TB and DS-TB isolates was obtained from MGIT cultures. The mean size for the MDR-TB isolates was 56,150 bp (Standard Deviation (SD)=6171) (MGIT) and 30,439 bp (SD=13,890) (7H11), whilst for the DS-TB isolates it was 54,776 bp (SD=5028) (MGIT) and 18,618 bp (SD=7134) (7H11). There were no significant differences between the means of DNA sizes for isolates grown in MGITs or on 7H11 for either MDR-TB or DS-TB isolates. For the MDR-TB isolates, when cultured in MGIT, the DNA size increased over time, whereas on 7H11, the DNA size peaked at week 3 (p=0.01, one-way ANOVA). For the DS-TB isolates, the increased over time in both the MGIT and 7H11 cultures.

DNA yield and DNA size results from the precipitation and spin-column CTAB extraction method were compared for MGIT and 7H11 cultures and stratified into resistance categories (Figure 2).

**Figure 2.**
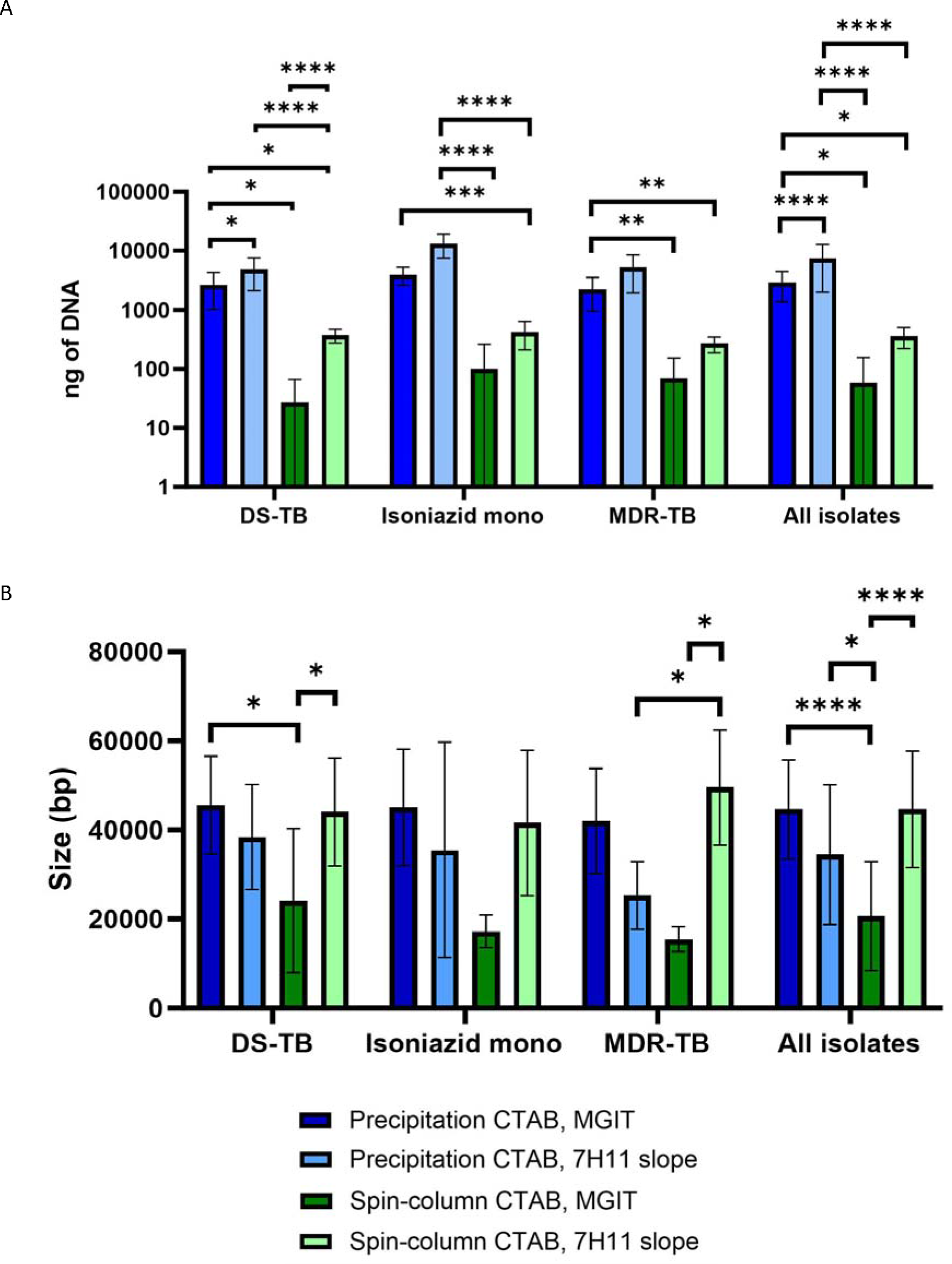
Precipitation and spin-column CTAB DNA extraction method comparison. A) DNA yield measured using Qubit dsDNA broad spectrum kit, B) Size (bp) measured using the TapeStation genomic DNA kit. Apart from spin-column CTAB from MGIT tubes, all other methods provided the minimum required yield, although precipitation based CTAB protocols provided higher yields. Spin-column CTAB from MGIT media also provided the smallest size. Significance was calculated using One-Way ANOVA and Tukey’s post ad hoc analysis.

We carried out MinION-based sequencing of selected samples. Firstly we looked at fragment size, as in our experience, sequencing coverage can be reduced both by fragments that are too short and too long. Long fragment lengths can especially inhibit read output for the mycobacteria. To do this we generated shorter DNA fragments (20 and 10 kb average size) which we compared with the original (50 kb) samples. A size of 20 kb resulted in the greatest number of reads, bases and N50 (Figure 3). The obtained N50 in sequencing was significantly higher with the 20 kb input DNA fragments compared to the other two sizes (p=0.01, one-way ANOVA). Although there was variation in the number of reads and bases obtained between the DNA sizes tested, the differences were not significant (p>0.05, one-way ANOVA).

**Figure 3.**
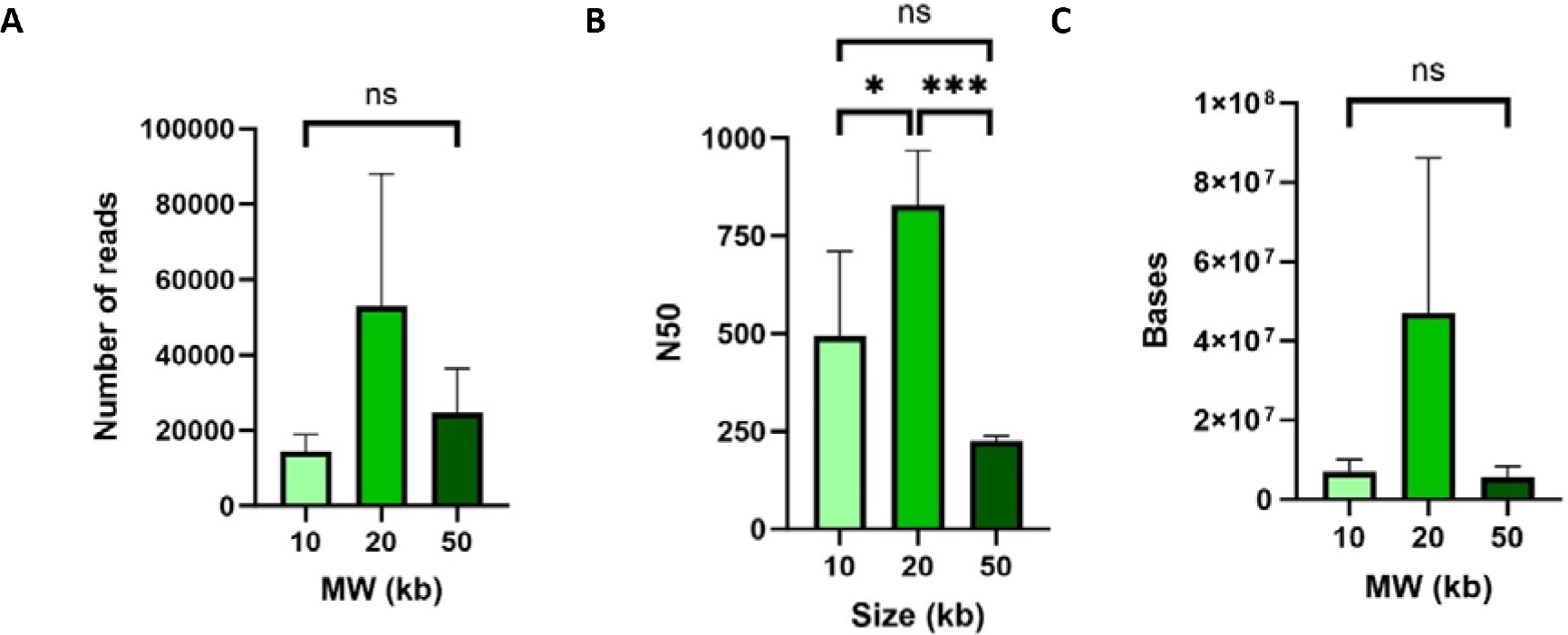
DNA shearing to 10 kb and 20 kb compared to 50 kb. A) number of reads (no significant difference) B) N50 (overall p=0.01) and C) total number of bases (no significant difference). One-Way ANOVA and Tukey’s multiple comparison test was applied.

We compared different library preparation kits, all three of which are rapid, transposase-based kits. SQK-RPB004 used a PCR amplification step resulting in fragments of around 4000 bp, the other two were different versions of kits that utilise a transposase that cuts native DNA to add the sequencing adapter, and has a major impact on fragment length. When 12 randomly selected isolates were barcoded and multiplexed and sequenced on one flow cell, the PCR-based SQK-RPB004 kit gave a greater mean read depth (mean=394x, SD=762) than the native kits, but the genome coverage was low (mean=67%, SD=19). Whilst the coverage was improved using the older native DNA-based kit (SQK-RBK004) (mean=80%, SD=38), the read depth was lower (mean=4x, SD=3). The newer native DNA-based kit (SQK-RBK110.96) provided an improved depth for both precipitation DNA extraction method (mean=13x, SD=9) and spin-column CTAB method (mean=49x, SD=36), and improved genome coverage (mean=84.6%, SD=31 and 99.8%, SD=2 respectively). Figure 4 highlights the quality scores for each of the kits. When 24 samples were run together, the mean depth was reduced to 18x (SD=25).

**Figure 4.**
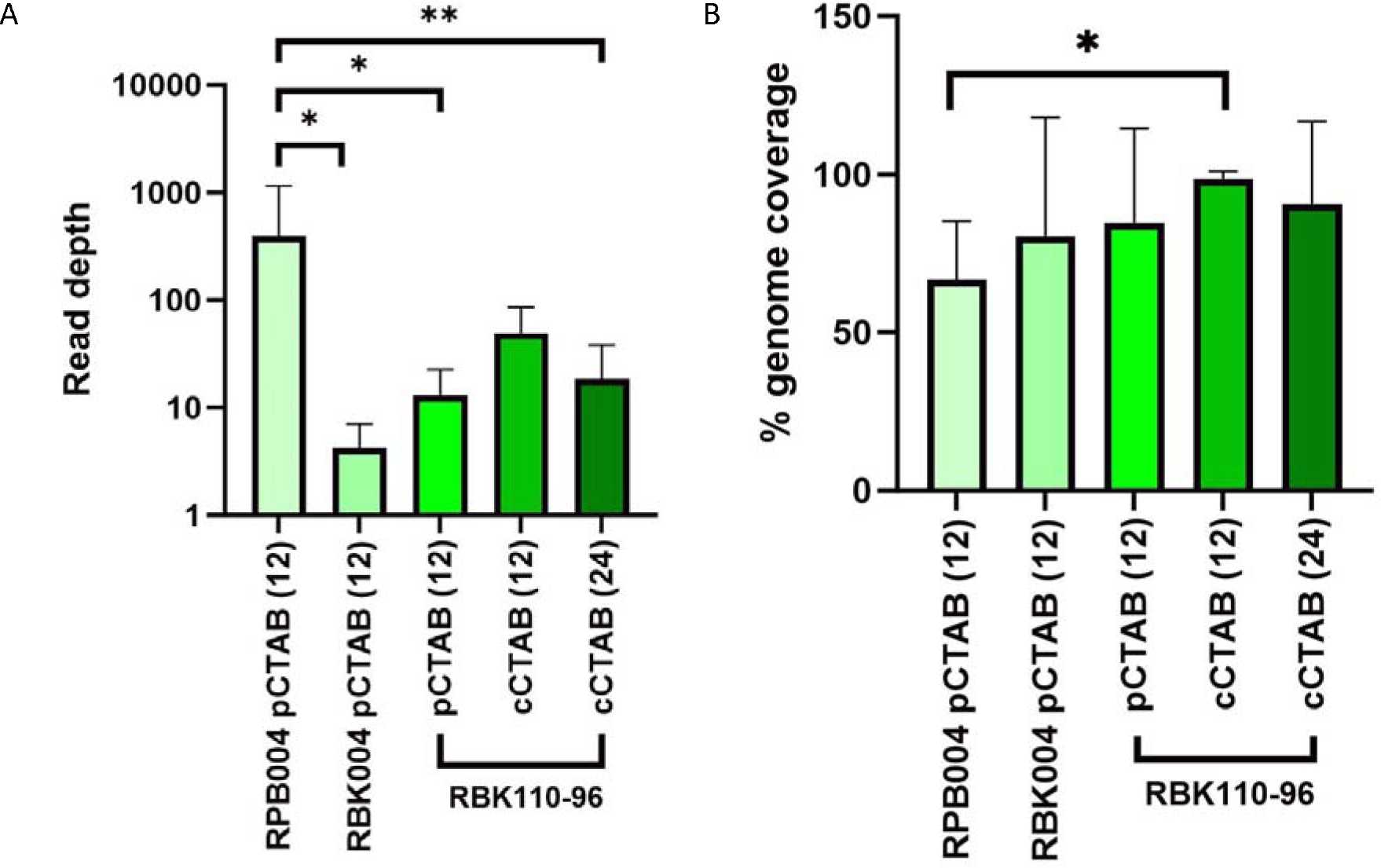
A) Mean read depth and B) genome coverage when comparing each of the three kits. Statistical significance calculated using One-Way ANOVA and Tukey’s multiple comparisons test. pCTAB = precipitation CTAB, cCTAB = spin column CTAB. Numbers in brackets denote the number of samples multiplexed per run.

When the assembled BAM files were viewed for rpoB, where most rifampicin-resistance SNPS map (as an example), the PCR-based SQK-RPB004 kit showed isolated, highly amplified regions and other regions with no coverage (see Figure 5). Native library preparation kits demonstrated a better coverage for the whole rpoB gene.

**Figure 5.**
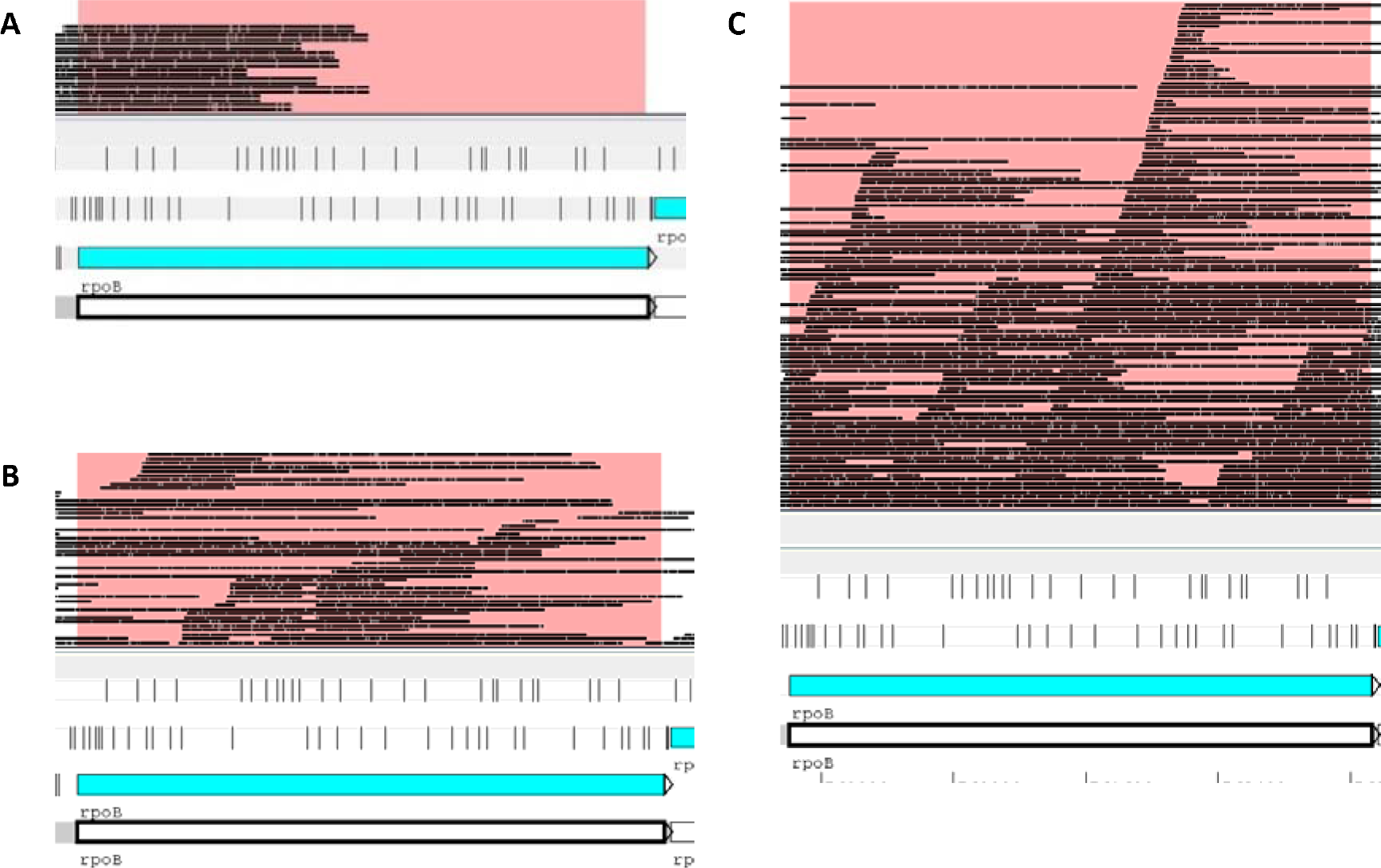
Comparison of BAM file read depth of rpoB for A) the PCR-based SQK-RPB004 kit, showing low coverage, B) SQK-RBK004 kit showing greater coverage and C) SQK-RBK110.96 resulted in the best coverage and depth, using Artemis.

Here we compared three different bioinformatic tools Fast basecalling was compared with high accuracy (HAC) and super high accuracy (SUP) basecalling. The mean number of .fast5 input files was 6.4 (SD=1.5) (Table 1). Whilst SUP basecalling was the slowest, it did provide significantly better quality (Q) scores.

**Table 1.** Comparison of basecalling algorithms. One-way ANOVA was used to test statistical significance. When Tukey’s multiple comparisons test was applied, there was a significant difference of p=<0·0001 between each of the basecalling algorithms for time, mean Q score and accuracy.

|  | Fast | HAC | SUP | One-way ANOVA significance |
| --- | --- | --- | --- | --- |
| Mean time (minutes) (SD) | 15 (5.9) | 205 (65.4) | >36 hours | $p < 0.0001$ |
| Mean Q score (SD) | 17.1 (0.2) | 19.7 (0.3) | 20.8 (0.3) | $p<0.0001$ |
| Mean accuracy (SD) | 98.02% (0.06) | 98.94% (0.05) | 99.13% (0.05) | $p<0.0001$ |

The DNA samples came from strains with known DR phenotypes and genotypes. Fast, HAC and SUP-basecalled versions of each of the files were analysed using TB-Profiler to compare resistance SNP and lineage identification (Table 2). All isolate’s median read depths were >40x. Compared with SUP basecalling, Fast basecalling agreed for 15/17 (88%) samples for lineage and all resistance SNPs. The sample for which Fast basecalling missed a SNP (streptomycin gidC 102de1G) had a depth of 47x. The sample for which Fast basecalling called lineage 3, and HAC and SUP called lineage 4.3, had a depth of 85x. HAC and SUP basecalling agreed for 17/17 (100%) samples. For the complete basecalling comparison dataset, see Supplementary materials S4.

**Table 2.** Concordance of different basecalling models, assuming greatest accuracy using SUP basecalling. HAC and SUP were 100% concordant, Fast basecalling called one lineage differently and missed one drug resistance SNP, compared to HAC and SUP basecalling. Isoniazid mono-resistance is defined by phenotypic DSTs.

|  | Average read depth | Fast vs SUP |  |  | HAC vs SUP |  |  |
| --- | --- | --- | --- | --- | --- | --- | --- |
|  |  | Lineage | Drug resistance | Total concordance | Lineage | Drug resistance | Total concordance |
| DS-TB | 90 | 7/8 (88%) | 8/8 (100%) | 7/8 (88%) | 8/8 (100%) | 8/8 (100%) | 8/8 (100%) |
| Isoniazid mono | 111 | 5/5 (100%) | 4/5 (80%) | 4/5 (80%) | 5/5 (100%) | 5/5 (100%) | 5/5 (100%) |
| MDR-TB | 72 | 4/4 (100%) | 4/4 (100%) | 4/4 (100%) | 4/4 (100%) | 4/4 (100%) | 4/4 (100%) |
| Total isolates | 85 | 16/17 (94%) | 16/17 (94%) | 15/17 (88%) | 17/17 (100%) | 17/17 (100%) | 17/17 (100%) |

Using phylogenetic DSTs as a guide, HAC-basecalled data were put through TB-Profiler and Mykrobe. Mykrobe called a mixed population of lineage 3 and 4.6.2.2 for the sample where Fast, HAC and SUP disagreed in TB-Profiler (Table 3). The complete dataset can be found in Supplementary materials S5.

**Table 3.** Summary comparison of the TB sequencing data analysis software Mykrobe, as compared against TB-Profiler.

| **Mykrobe vs TB-Profiler** | **Lineage** | **Drug resistance** | **Total concordance** |
| --- | --- | --- | --- |
| DS-TB | 7/8 (88%) | 7/8 (88%) | 7/8 (88%) |
| Isoniazid mono-resistance | 5/5 (100%) | 5/5 (100%) | 5/5 (100%) |
| MDR-TB | 4/4 (100%) | 1/4 (25%) | 1/4 (25%) |
| Total isolates | 16/17 (94%) | 13/17 (76%) | 13/17 (76%) |

HAC-basecalled consensus sequence fastq files were compared with the equivalent Illumina fastq files for each isolate, to identify lineage and resistance SNP concordance using TB-Profiler. ONT data was concordant with phenotypic DSTs in 12/17 (71%) samples, taking into account phenotypic DST testing only included rifampicin, isoniazid, ethambutol and streptomycin, and pyrazinamide for MDR samples. ONT picked up SNPs in one DS-TB sample, one isoniazid mono-resistant samples two MDR-TB samples which phenotypic DSTs concluded were sensitive. Both ONT and Illumina showed two phenotypically pyrazinamide-resistant isolates as sensitive. The full list of phenotypic and genotypic drug resistances found, including full SNP calls are in Supplementary materials S6.

The ONT pipeline was overall 94% (16/17) concordant (16/17 for lineage and 17/17 for all resistance SNPs) with Illumina data. For lineage, one DS-TB sample appeared to be a mixed population of lineage 3 (the only one picked up by Illumina) and 4.6 (picked up by HAC and SUP-basecalling in the ONT pipeline, but not Fast). An overview of these results is in Figure 6.

**Figure 6.**
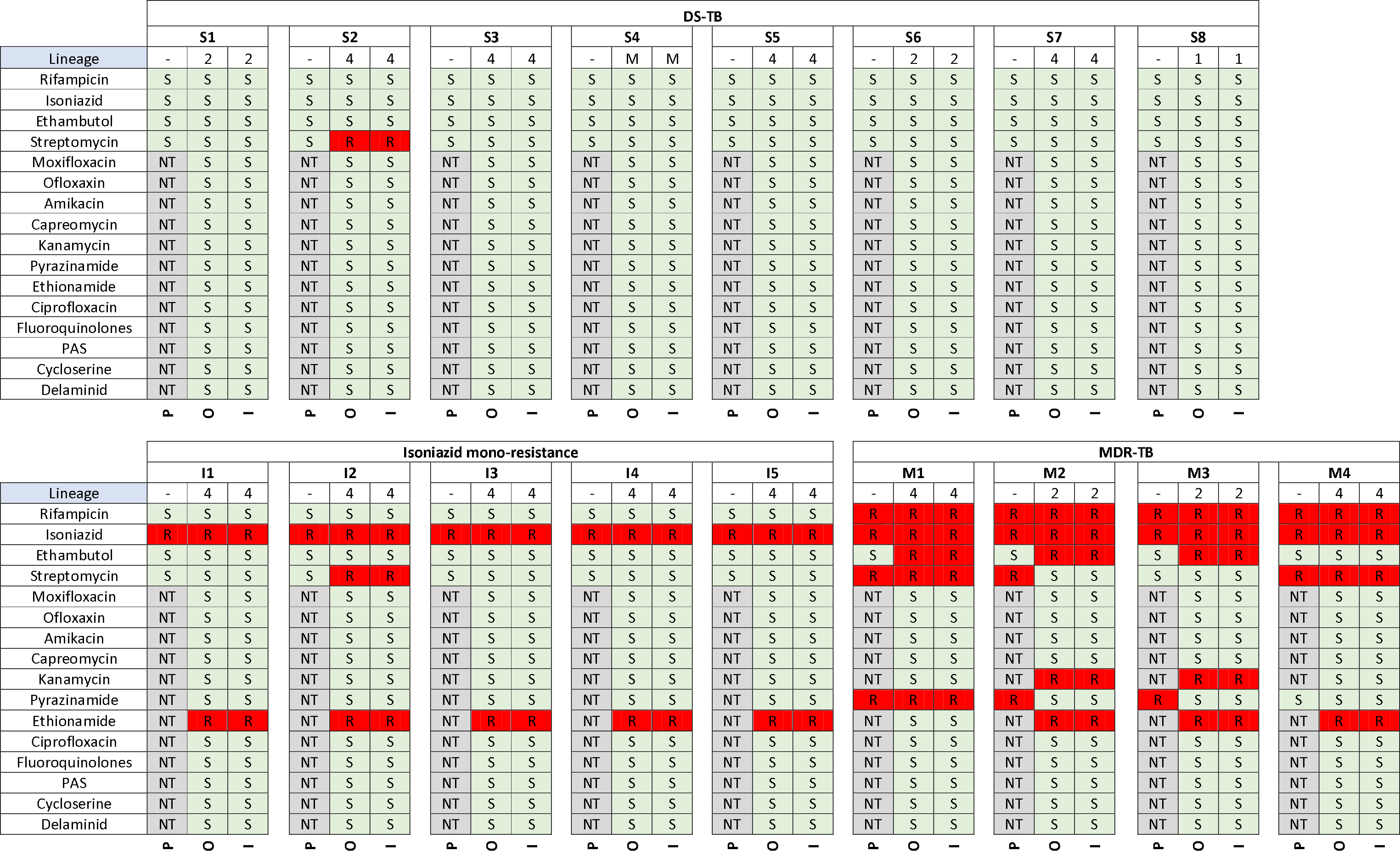
Drug resistance and lineage profile concordance resulting from phenotypic DSTs, ONT and Illumina sequencing. P = phenotypic DST results, O = Oxford Nanopore results, I = Illumina results, M = mixed (lineage 3 and 4), S = sensitive, R = resistant, NT = not tested.

Table 4 outlines each component analysed, which tested component performed the best for each category, and the overall recommended pipeline options.

**Table 4.**
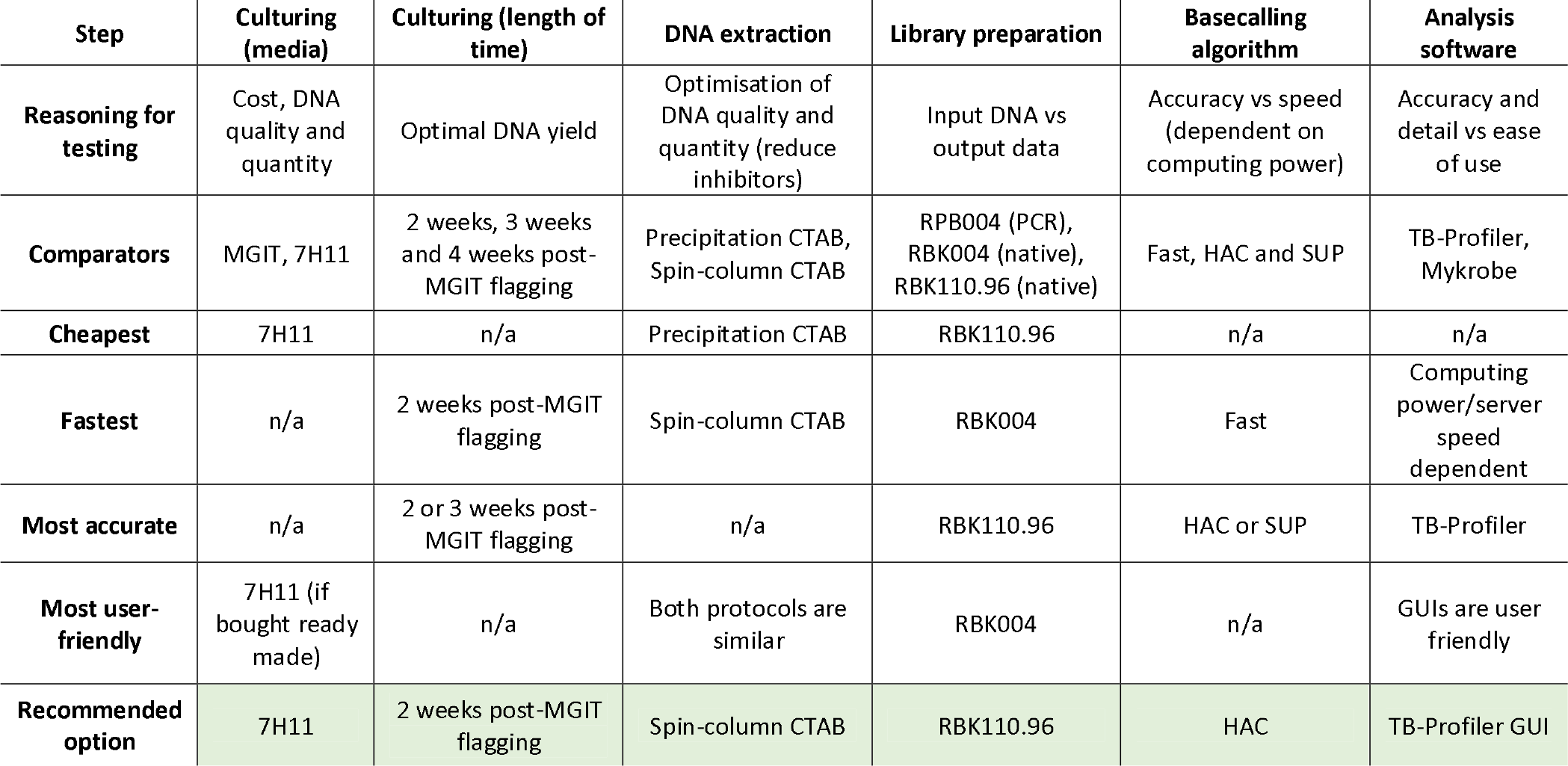
Table of pipeline steps and overall recommended choice for each section of the pipeline.

| Step | Culturing (media) | Culturing (length of time) | DNA extraction | Library preparation | Basecalling algorithm | Analysis software |
| --- | --- | --- | --- | --- | --- | --- |
| **Reasoning for testing** | Cost, DNA quality and quantity | Optimal DNA yield | Optimisation of DNA quality and quantity (reduce inhibitors) | Input DNA vs output data | Accuracy vs speed (dependent on computing power) | Accuracy and detail vs ease of use |
| **Comparators** | MGIT, 7H11 | 2 weeks, 3 weeks and 4 weeks post-MGIT flagging | Precipitation CTAB, Spin-column CTAB | RPB004 (PCR), RBK004 (native), RBK110.96 (native) | Fast, HAC and SUP | TB-Profiler, Mykrobe |
| **Cheapest** | 7H11 | n/a | Precipitation CTAB | RBK110.96 | n/a | n/a |
| **Fastest** | n/a | 2 weeks post-MGIT flagging | Spin-column CTAB | RBK004 | Fast | Computing power/server speed dependent |
| **Most accurate** | n/a | 2 or 3 weeks post-MGIT flagging | n/a | RBK110.96 | HAC or SUP | TB-Profiler |
| **Most user-friendly** | 7H11 (if bought ready made) | n/a | Both protocols are similar | RBK004 | n/a | GUIs are user friendly |
| **Recommended option** | 7H11 | 2 weeks post-MGIT flagging | Spin-column CTAB | RBK110.96 | HAC | TB-Profiler GUI |

## Discussion

We conclude that a WGS pipeline using ONT technology is feasible for the identification of SNPs that determine M. tuberculosis drug resistance and lineage and is suitable for our aim to create a pragmatic pipeline for use in LMICs. Taking into consideration the data presented here, we propose the following pipeline (Figure 7) as the optimized route for TB WGS.

**Figure 7.**
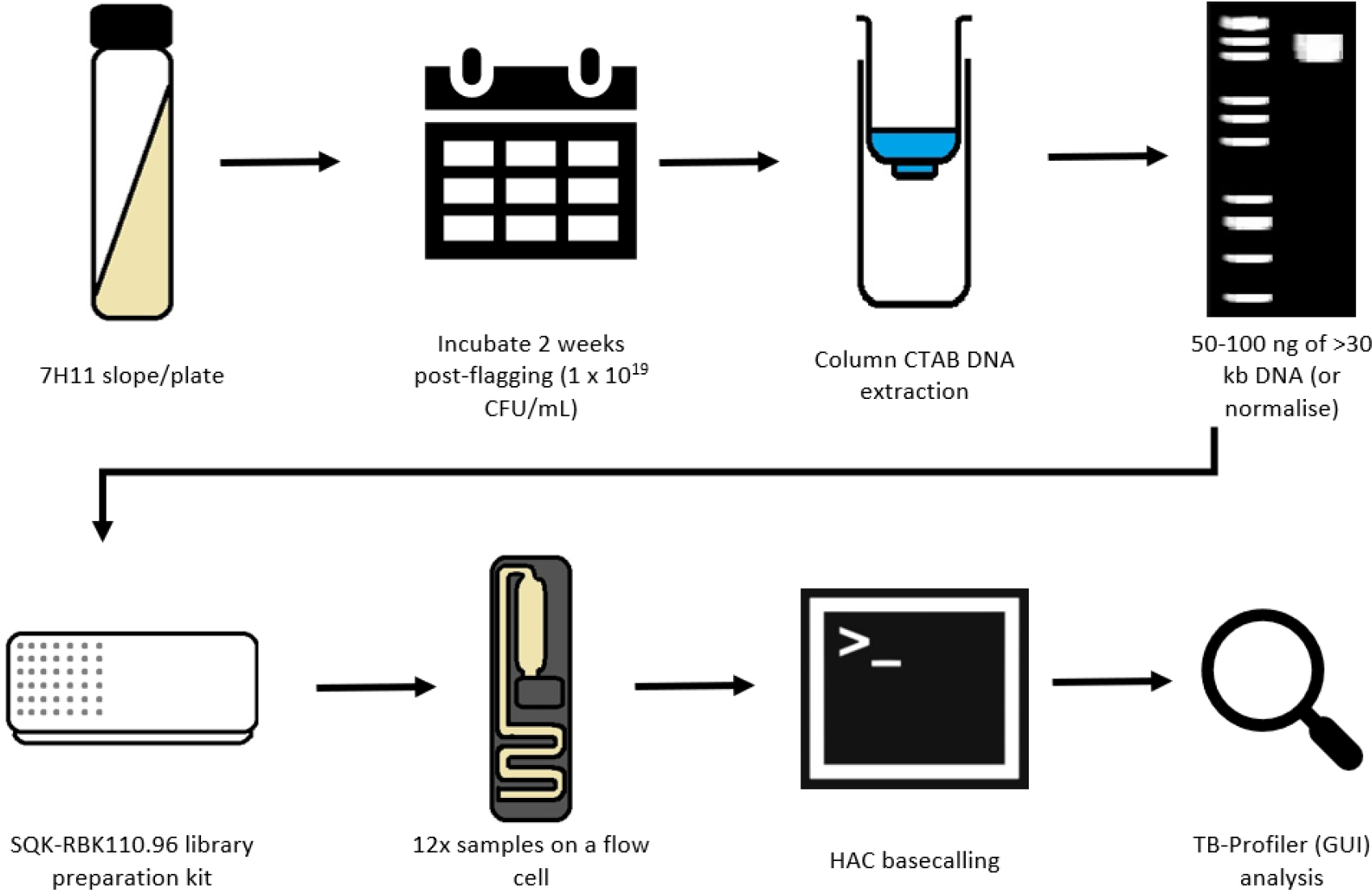
The recommended ONT WGS TB pipeline, including culturing conditions, DNA extraction methodology, optimal DNA input library preparation kit, basecalling and data analysis. Images taken from Windows 10 Education and ShutterStock.

In our study, the ONT pipeline showed the same accuracy as Illumina but showed less concordance with phenotypic DSTs. This is likely due to the complex relationship between genotypic and phenotypic resistance. All phenotypic and genotypic discordances in this study were seen in three drugs; streptomycin, ethambutol and pyrazinamide, each of which are known to have multifaceted resistance mechanisms, which are not fully understood, thus further work is needed (46).

We identified isolates with phenotypic sensitivity and genotypic resistance, which may be a result of subpopulations and heteroresistance, which is a growing concern. When phenotypic resistance and genotypic sensitivity was seen, this may have been a result of the specific resistance SNPs causing genotypic resistance not being included in the analysis software databases. Ultimately, the standard will be which drugs are clinically relevant, and whilst phenotypic DSTs are the current benchmark, with the accumulation of genotypic resistance knowledge, this may shift to a molecular benchmark in the future.

Whilst a reduced fragment size did improve sequencing read yield, the Covaris g-TUBEs added a significant cost to the pipeline. We found that extracting from 7H11 plates in conjunction with spin- column CTAB DNA extraction, produced a fragment length of <30 kb. Thus, we recommend that g- TUBEs could be used as an optional extra within the pipeline, if different media and extraction methodologies are utilised.

Whilst the precipitation CTAB method does provide adequate DNA size and yield for use with the new native ONT library preparation kit (SQK-RBK110.96), lower numbers of reads and bases were generally achieved compared with spin-column CTAB extraction, although the latter produces lower DNA yields. We hypothesise that the co-precipitation of inhibitors, possibly from the mycobacterial cell wall, may be interfering with the binding of the transposase complex used in native library preparation kits, so that less DNA is adapted and therefore is unable to be sequenced.

The cost of WGS has decreased greatly in the last few years, making it more accessible for resource constrained settings. Whilst the cost per sample may be lower using Illumina due to the higher throughput, if the samples have to be sent away due to a lack of local facilities, extra costs are likely to be incurred. If batching is used to reduce costs, then this can increase the time to results in smaller laboratories with fewer samples. Costs in this pipeline could be reduced by substituting products or equipment, although automated systems and high-speed computers may be beneficial in the long term.

The bottleneck of this pipeline is likely to be basecalling, unless high-computing power set ups are available. The time from culture to drug resistance and lineage results could be within four weeks if the appropriate human resources and computers are available, with 2-3 weeks of culturing and a week for extraction, quantification, sequencing and analysis. Whilst this is not as fast as other methods such as GeneXpert or line probe assays which are commonly available in local regional laboratories, it is comparable with phenotypic DSTs. In terms of the ease-of-use of this pipeline, if users opt to use Guppy within the MinKNOW GUI, minimal CLI knowledge is required, meaning less training would be required for routine laboratory staff, and should therefore be faster to implement and more widely usable.

Some complications were encountered in this study, and in sequencing of the mycobacteria in general. This pipeline requires cultured isolates, and thus the use of a biosafety level 3 (BSL3) laboratory, which are not widely available. The possibility of utilising another site’s BSL3 facilities to grow cultures, heat killing and then transferring samples to the sequencing laboratory is an alternative route but may increase the time to results.

With a greater understanding of genomic drug resistances, plus ever-evolving bioinformatics software, the accuracy TB diagnostics will improve. To increase the speed of time to results, sequencing direct from sputum would cut out culturing time, but is yet to be standard practice in routine diagnostics. Targeted kits, such as the Illumina and GenoScreen Deeplex® Myc-TB Combo Kit are an alternative to WGS but provide less information (47). An ONT targeted kit for DR-TB has recently been approved by the WHO (48).

Whilst the necessity of culture in this pipeline does mean time to results is in weeks not minutes or days, the resolution of resistance and lineage detail obtained from ONT WGS sequencing is greater than that obtained from standard diagnostic tests such as GeneXpert, line probe assays or phenotypic DSTs, and is comparable to Illumina sequencing. There is a need for genotypic drug resistance data, not only for routine diagnostics, but also for geographical drug resistance profiling and surveillance to aid correct prescribing, and it is important that data is collected as broadly as possible (9). The ability of smaller, localised laboratories to participate in this is vital and the optimised ONT pipeline presented here will help to facilitate WGS roll out within these sites.

## Data sharing statement

All genetic fasta data obtained in this study is available under the Project accession number PRJEB68143 on the European Nucleotide Archive upon publication. The full manual, including further information and tutorial videos, relating to this protocol is available on the PANDORA-ID-NET Global Health Network hub pages: https://pandora.tghn.org/sequencing/tuberculosis-sequencing-pipeline/. All data and additional material is Open Access.

## Abbreviations

|  |  |
| --- | --- |
| CTAB | Cethyl trimethyl ammonium bromide |
| CLI | Command line interface |
| CPU | Central processing unit |
| DR-TB | Drug resistant tuberculosis |
| DS-TB | Drug sensitive tuberculosis |
| DST | Drug sensitivity test |
| GPU | Graphics processing unit |
| GUI | Graphical user interface |
| HAC | High accuracy (basecalling) |
| LMIC | Low- and middle-income country |
| MDR | Multi-drug resistant |
| ONT | Oxford Nanopore Technologies |
| QC | Quality control |
| RoC | Republic of Congo |
| RR | Rifampicin-resistant |
| SD | Standard deviation |
| SUP | Super-high accuracy (basecalling) |
| TB | Tuberculosis |
| WGS | Whole genome sequencing |
| XDR | Extensively-drug resistant |

