## Supplementary materials for "A pragmatic pipeline for drug resistance identification in *Mycobacterium tuberculosis* using whole genome sequencing"

**Contents**

**S1 – Full column CTAB protocol** 1

**S2 - Computer specifications for the computer used in this project** 1

**S3 - List of uploaded fastq files** 1

**S4 - Full basecalling comparison table** 3

**S5 - Full tuberculosis data analysis software comparison table** 5

**S6 - Full ONT vs Illumina concordance table** 7

### S1 – Full column CTAB protocol

500 µL of liquid culture or a loopful of solid culture was resuspended in 500 µL PBS in a 2 mL MPBio tube, heat killed, washed twice with PBS and resuspended in 100 µL TE buffer (10mM Tris pH8 and 1mM EDTA). 2 x 4 mm glass beads were added and the samples were vortexed for 45 seconds. 10 µL lysozyme (10 mg/mL) was added and the samples then incubated at 37°C for 15 minutes at 800 RPM. The samples were then transferred into 1.5 mL Eppendorf DNA LoBind tubes and 10 µL proteinase K and 3 µL RNase A added, provided with the NEB Monarch Genomic Purification Kit (T3010S, New England Biolabs). This was vortexed, 200 µL CTAB buffer (2% CTAB, 40mM EDTA, 1.4M NaCl and 100mM Tris pH8.0) added and vortexed again. The samples were incubated at 56°C for 30 minutes at 1000 RPM, then 100 µL 5M NaCL added. The tubes were centrifuged at 17,000 x *g* for 5 minutes and 200 µL of the supernatant retained, avoiding the pellet.

The Monarch Genomic Purification Kit manufacturer instructions were then followed, with these alterations: 400 µL gDNA binding buffer was added to the supernatant and vortexed for 10 seconds. The lysate/binding buffer mix was transferred to a gDNA purification column, centrifuged at 1,000 x g for 3 minutes and then maximum speed for 1 minute to clear the membrane. 500 µL gDNA wash buffer was added and centrifuged for 1 minute at maximum speed and then repeated. The gDNA purification column was then placed in a DNase-free 2.5 mL microfuge tube and 50-100 µL preheated 60°C gDNA elution buffer added. The tubes were incubated at room temperature for 1 minute and then centrifuged at maximum speed for 1 minute to elute the gDNA. DNA was eluted into 50 µL molecular grade water.

### S2 - Computer specifications for the computer used in this project

| Component Required | Specification |
|---|---|
| Operating system | Windows 10 |
| Memory/RAM | 32 GB RAM |
| CPU | Intel(R) Core(TM) i7-10700 CPU @ 2.90GHz 2.90 GHz |
| Storage | 1 TB internal SSD |
| Ports | USB 3.0 |

### S3 - List of uploaded fastq files

| Isolate name | Sequencing platform | Sample Accession number | Experiment Accession number |
|---|---|---|---|
| TB_sensitive_1 | Oxford Nanopore Technologies | ERS16613404 | ERX11656854 |
| TB_sensitive_2 | Oxford Nanopore Technologies | ERS16613405 | ERX11656855 |
| TB_sensitive_3 | Oxford Nanopore Technologies | ERS16613406 | ERX11656856 |
| TB_sensitive_4 | Oxford Nanopore Technologies | ERS16613407 | ERX11656857 |
| TB_sensitive_5 | Oxford Nanopore Technologies | ERS16613408 | ERX11656858 |
| TB_sensitive_6 | Oxford Nanopore Technologies | ERS16613409 | ERX11656859 |
| TB_sensitive_7 | Oxford Nanopore Technologies | ERS16613410 | ERX11656860 |
| TB_sensitive_8 | Oxford Nanopore Technologies | ERS16613411 | ERX11656861 |
| TB_isoniazid_mono_1 | Oxford Nanopore Technologies | ERS16613412 | ERX11656845 |
| TB_isoniazid_mono_2 | Oxford Nanopore Technologies | ERS16613413 | ERX11656846 |

| | | | |
|---|---|---|---|
| TB_isoniazid_mono_3 | Oxford Nanopore Technologies | ERS16613415 | ERX11656847 |
| TB_isoniazid_mono_4 | Oxford Nanopore Technologies | ERS16613416 | ERX11656848 |
| TB_isoniazid_mono_5 | Oxford Nanopore Technologies | ERS16613417 | ERX11656849 |
| TB_MDR_1 | Oxford Nanopore Technologies | ERS16613418 | ERX11656850 |
| TB_MDR_2 | Oxford Nanopore Technologies | ERS16613419 | ERX11656851 |
| TB_MDR_3 | Oxford Nanopore Technologies | ERS16613420 | ERX11656852 |
| TB_MDR_4 | Oxford Nanopore Technologies | ERS16613421 | ERX11656853 |
| TB_sensitive_1_1 | Illumina | ERS16690010 | ERX11656882 |
| TB_sensitive_1_2 | Illumina | ERS16690011 | ERX11656883 |
| TB_sensitive_2_1 | Illumina | ERS16690012 | ERX11656884 |
| TB_sensitive_2_2 | Illumina | ERS16690013 | ERX11656885 |
| TB_sensitive_3_1 | Illumina | ERS16690014 | ERX11656886 |
| TB_sensitive_3_2 | Illumina | ERS16690015 | ERX11656887 |
| TB_sensitive_4_1 | Illumina | ERS16690016 | ERX11656888 |
| TB_sensitive_4_2 | Illumina | ERS16690017 | ERX11656889 |
| TB_sensitive_5_1 | Illumina | ERS16690018 | ERX11656890 |
| TB_sensitive_5_2 | Illumina | ERS16690019 | ERX11656891 |
| TB_sensitive_6_1 | Illumina | ERS16690020 | ERX11656892 |
| TB_sensitive_6_2 | Illumina | ERS16690021 | ERX11656893 |
| TB_sensitive_7_1 | Illumina | ERS16690022 | ERX11656894 |
| TB_sensitive_7_2 | Illumina | ERS16690023 | ERX11656895 |
| TB_sensitive_8_1 | Illumina | ERS16690024 | ERX11656896 |
| TB_sensitive_8_2 | Illumina | ERS16690025 | ERX11656897 |
| TB_isoniazid_mono_1_1 | Illumina | ERS16690026 | ERX11656864 |
| TB_isoniazid_mono_1_2 | Illumina | ERS16690027 | ERX11656865 |
| TB_isoniazid_mono_2_1 | Illumina | ERS16690028 | ERX11656866 |
| TB_isoniazid_mono_2_2 | Illumina | ERS16690029 | ERX11656867 |
| TB_isoniazid_mono_3_1 | Illumina | ERS16690030 | ERX11656868 |
| TB_isoniazid_mono_3_2 | Illumina | ERS16690031 | ERX11656869 |
| TB_isoniazid_mono_4_1 | Illumina | ERS16690032 | ERX11656870 |
| TB_isoniazid_mono_4_2 | Illumina | ERS16690033 | ERX11656871 |
| TB_isoniazid_mono_5_1 | Illumina | ERS16690034 | ERX11656872 |
| TB_isoniazid_mono_5_2 | Illumina | ERS16690035 | ERX11656873 |
| TB_MDR_1_1 | Illumina | ERS16690036 | ERX11656874 |
| TB_MDR_1_2 | Illumina | ERS16690037 | ERX11656875 |
| TB_MDR_2_1 | Illumina | ERS16690038 | ERX11656876 |
| TB_MDR_2_2 | Illumina | ERS16690039 | ERX11656877 |
| TB_MDR_3_1 | Illumina | ERS16690040 | ERX11656878 |
| TB_MDR_3_2 | Illumina | ERS16690041 | ERX11656879 |
| TB_MDR_4_1 | Illumina | ERS16690042 | ERX11656880 |
| TB_MDR_4_2 | Illumina | ERS16690043 | ERX11656881 |

### S4 - Full basecalling comparison table

| | Sample | Basecaller | Lineage | Lineage name | Rifampicin | Isoniazid | Ethambutol | Streptomycin | Moxifloxacin | Ofloxacin | Amikacin | Capreomycin | Kanamycin | Pyrazinamide | Ethionamide | Ciprofloxacin | Fluoroquinolones | PAS | Cycloserine | Delaminid |
|---|---|---|---|---|---|---|---|---|---|---|---|---|---|---|---|---|---|---|---|---|
| Sensitive | S1 | F | 2.2.1 | Beijing | Se | Se | Se | Se | Se | Se | Se | Se | Se | Se | Se | Se | Se | Se | Se | Se |
| | | H | 2.2.1 | Beijing | Se | Se | Se | Se | Se | Se | Se | Se | Se | Se | Se | Se | Se | Se | Se | Se |
| | | S | 2.2.1 | Beijing | Se | Se | Se | Se | Se | Se | Se | Se | Se | Se | Se | Se | Se | Se | Se | Se |
| | S2 | F | 4.1.2.1 | None | Se | Se | Se | rpsL p.Lys43Arg | Se | Se | Se | Se | Se | Se | Se | Se | Se | Se | Se | Se |
| | | H | 4.1.2.1 | T1 | Se | Se | Se | rpsL p.Lys43Arg | Se | Se | Se | Se | Se | Se | Se | Se | Se | Se | Se | Se |
| | | S | 4.1.2.1 | T1 | Se | Se | Se | rpsL p.Lys43Arg | Se | Se | Se | Se | Se | Se | Se | Se | Se | Se | Se | Se |
| | S3 | F | 4.1.1.1 | X2 | Se | Se | Se | Se | Se | Se | Se | Se | Se | Se | Se | Se | Se | Se | Se | Se |
| | | H | 4.1.1.1 | X2 | Se | Se | Se | Se | Se | Se | Se | Se | Se | Se | Se | Se | Se | Se | Se | Se |
| | | S | 4.1.1.1 | X2 | Se | Se | Se | Se | Se | Se | Se | Se | Se | Se | Se | Se | Se | Se | Se | Se |
| | S4 | F | 3 | CAS | Se | Se | Se | Se | Se | Se | Se | Se | Se | Se | Se | Se | Se | Se | Se | Se |
| | | H | 4.6 | Manu2 | Se | Se | Se | Se | Se | Se | Se | Se | Se | Se | Se | Se | Se | Se | Se | Se |
| | | S | 4.6 | Manu2 | Se | Se | Se | Se | Se | Se | Se | Se | Se | Se | Se | Se | Se | Se | Se | Se |
| | S5 | F | 4.3.4 | None | Se | Se | Se | Se | Se | Se | Se | Se | Se | Se | Se | Se | Se | Se | Se | Se |
| | | H | 4.3.4.2 | LAM9 | Se | Se | Se | Se | Se | Se | Se | Se | Se | Se | Se | Se | Se | Se | Se | Se |
| | | S | 4.3.4.2 | LAM9 | Se | Se | Se | Se | Se | Se | Se | Se | Se | Se | Se | Se | Se | Se | Se | Se |
| | S6 | F | 2.2.1 | Beijing | Se | Se | Se | Se | Se | Se | Se | Se | Se | Se | Se | Se | Se | Se | Se | Se |
| | | H | 2.2.1 | Beijing | Se | Se | Se | Se | Se | Se | Se | Se | Se | Se | Se | Se | Se | Se | Se | Se |
| | | S | 2.2.1 | Beijing | Se | Se | Se | Se | Se | Se | Se | Se | Se | Se | Se | Se | Se | Se | Se | Se |
| | S7 | F | 4.1.3 | None | Se | Se | Se | Se | Se | Se | Se | Se | Se | Se | Se | Se | Se | Se | Se | Se |

| | | | | | | | | | | | | | | | | | | | | |
|---|---|---|---|---|---|---|---|---|---|---|---|---|---|---|---|---|---|---|---|---|
| | | H | 4.1.3 | T1 | Se | Se | Se | Se | Se | Se | Se | Se | Se | Se | Se | Se | Se | Se | Se | Se |
| | | S | 4.1.3 | T1 | Se | Se | Se | Se | Se | Se | Se | Se | Se | Se | Se | Se | Se | Se | Se | Se |
| | S8 | F | 1.2.1.2.1 | EAI2-nonthaburi | Se | Se | Se | Se | Se | Se | Se | Se | Se | Se | Se | Se | Se | Se | Se | Se |
| | | H | 1.2.1.2.1 | EAI2-nonthaburi | Se | Se | Se | Se | Se | Se | Se | Se | Se | Se | Se | Se | Se | Se | Se | Se |
| | | S | 1.2.1.2.1 | EAI2-nonthaburi | Se | Se | Se | Se | Se | Se | Se | Se | Se | Se | Se | Se | Se | Se | Se | Se |
| Isoniazid mono-resistant | I1 | F | 4.6.2.2 | None | Se | fabG1 c.-15C>T | Se | Se | Se | Se | Se | Se | Se | Se | fabG1 c.-15C>T | Se | Se | Se | Se | Se |
| | | H | 4.6.2.2 | Cameroon | Se | fabG1 c.-15C>T | Se | Se | Se | Se | Se | Se | Se | Se | fabG1 c.-15C>T | Se | Se | Se | Se | Se |
| | | S | 4.6.2.3 | Cameroon | Se | fabG1 c.-15C>T | Se | Se | Se | Se | Se | Se | Se | Se | fabG1 c.-15C>T | Se | Se | Se | Se | Se |
| | I2 | F | 4.6.1.2 | None | Se | fabG1 c.-15C>T, inhA p.Ile194Thr | Se | Se | Se | Se | Se | Se | Se | Se | fabG1 c.-15C>T, inhA p.Ile194Thr | Se | Se | Se | Se | Se |
| | | H | 4.6.1.2 | X1 | Se | fabG1 c.-15C>T, inhA p.Ile194Thr | Se | gid c.102de1G | Se | Se | Se | Se | Se | Se | fabG1 c.-15C>T, inhA p.Ile194Thr | Se | Se | Se | Se | Se |
| | | S | 4.6.1.2 | X1 | Se | fabG1 c.-15C>T, inhA p.Ile194Thr | Se | gid c.102de1G | Se | Se | Se | Se | Se | Se | fabG1 c.-15C>T, inhA p.Ile194Thr | Se | Se | Se | Se | Se |
| | I3 | F | 4.6.2.2 | Cameroon | Se | fabG1 c.-15C>T | Se | Se | Se | Se | Se | Se | Se | Se | fabG1 c.-15C>T | Se | Se | Se | Se | Se |
| | | H | 4.6.2.2 | Cameroon | Se | fabG1 c.-15C>T | Se | Se | Se | Se | Se | Se | Se | Se | fabG1 c.-15C>T | Se | Se | Se | Se | Se |
| | | S | 4.6.2.2 | Cameroon | Se | fabG1 c.-15C>T | Se | Se | Se | Se | Se | Se | Se | Se | fabG1 c.-15C>T | Se | Se | Se | Se | Se |
| | I4 | F | 4.6.2.2 | Cameroon | Se | fabG1 c.-15C>T | Se | Se | Se | Se | Se | Se | Se | Se | fabG1 c.-15C>T | Se | Se | Se | Se | Se |
| | | H | 4.6.2.2 | Cameroon | Se | fabG1 c.-15C>T | Se | Se | Se | Se | Se | Se | Se | Se | fabG1 c.-15C>T | Se | Se | Se | Se | Se |
| | | S | 4.6.2.2 | Cameroon | Se | fabG1 c.-15C>T | Se | Se | Se | Se | Se | Se | Se | Se | fabG1 c.-15C>T | Se | Se | Se | Se | Se |
| | I5 | Fast | 4.6.2.2 | Cameroon | Se | fabG1 c.-15C>T | Se | Se | Se | Se | Se | Se | Se | Se | fabG1 c.-15C>T | Se | Se | Se | Se | Se |
| | | HAC | 4.6.2.2 | Cameroon | Se | fabG1 c.-15C>T | Se | Se | Se | Se | Se | Se | Se | Se | fabG1 c.-15C>T | Se | Se | Se | Se | Se |
| | | SUP | 4.6.2.2 | Cameroon | Se | fabG1 c.-15C>T | Se | Se | Se | Se | Se | Se | Se | Se | fabG1 c.-15C>T | Se | Se | Se | Se | Se |
| MDR | M1 | F | 4.1.2.1 | None | rpoB p.Ser450Leu | katG p.Ser315Thr | embB p.Met306Ile | rpsL p.Lys43Arg | Se | Se | Se | Se | Se | pncA p.Leu85Pro | Se | Se | Se | Se | Se | Se |
| | | H | 4.1.2.1 | H1 | rpoB p.Ser450Leu | katG p.Ser315Thr | embB p.Met306Ile | rpsL p.Lys43Arg | Se | Se | Se | Se | Se | pncA p.Leu85Pro | Se | Se | Se | Se | Se | Se |
| | | S | 4.1.2.1 | H1 | rpoB p.Ser450Leu | katG p.Ser315Thr | embB p.Met306Ile | rpsL p.Lys43Arg | Se | Se | Se | Se | Se | pncA p.Leu85Pro | Se | Se | Se | Se | Se | Se |

| | | | | | | | | | | | | | | | | | | | | |
|---|---|---|---|---|---|---|---|---|---|---|---|---|---|---|---|---|---|---|---|---|
| | M2 | F | 2.2.1 | Beijing | rpoB p.Ser450Leu, rpoB p.Glu761Asp | katG p.Ser315Thr | embB p.Asp354Ala | Se | Se | Se | Se | Se | eis c.-37G>T | Se | ethA c.-7T>C | Se | Se | Se | Se | Se |
| | | H | 2.2.1 | Beijing | rpoB p.Ser450Leu, rpoB p.Glu761Asp | katG p.Ser315Thr | embB p.Asp354Ala | Se | Se | Se | Se | Se | eis c.-37G>T | Se | ethA c.-7T>C | Se | Se | Se | Se | Se |
| | | S | 2.2.1 | Beijing | rpoB p.Ser450Leu, rpoB p.Glu761Asp | katG p.Ser315Thr | embB p.Asp354Ala | Se | Se | Se | Se | Se | eis c.-37G>T | Se | ethA c.-7T>C | Se | Se | Se | Se | Se |
| | M3 | F | 2.2.1 | Beijing | rpoB p.Ser450Leu, rpoB p.Glu761Asp | katG p.Ser315Thr | embB p.Asp354Ala | Se | Se | Se | Se | Se | eis c.-37G>T | Se | ethA c.-7T>C | Se | Se | Se | Se | Se |
| | | H | 2.2.1 | Beijing | rpoB p.Ser450Leu, rpoB p.Glu761Asp | katG p.Ser315Thr | embB p.Asp354Ala | Se | Se | Se | Se | Se | eis c.-37G>T | Se | ethA c.-7T>C | Se | Se | Se | Se | Se |
| | | S | 2.2.1 | Beijing | rpoB p.Ser450Leu, rpoB p.Glu761Asp | katG p.Ser315Thr | embB p.Asp354Ala | Se | Se | Se | Se | Se | eis c.-37G>T | Se | ethA c.-7T>C | Se | Se | Se | Se | Se |
| | M4 | F | 4.2.1 | Euro-American (TUR) | rpoB p.Ser450Leu, rpoC p.Asp485Asn | inhA c.-154G>A, katG p.Ser315Thr | Se | rpsL p.Lys88Arg | Se | Se | Se | Se | Se | Se | inhA c.-154G>A | Se | Se | Se | Se | Se |
| | | H | 4.2.1 | Ural-1 | rpoB p.Ser450Leu, rpoC p.Asp485Asn | inhA c.-154G>A, katG p.Ser315Thr | Se | rpsL p.Lys88Arg | Se | Se | Se | Se | Se | Se | inhA c.-154G>A | Se | Se | Se | Se | Se |
| | | S | 4.2.1 | Ural-1 | rpoB p.Ser450Leu, rpoC p.Asp485Asn | inhA c.-154G>A, katG p.Ser315Thr | Se | rpsL p.Lys88Arg | Se | Se | Se | Se | Se | Se | inhA c.-154G>A | Se | Se | Se | Se | Se |

F=fast basecalling, H= high accuracy basecalling, S= super high accuracy basecalling, Se=sensitive

**S5 - Full tuberculosis data analysis software comparison table**

| | Sample | Software | Lineage | Lineage name | Rifampicin | Isoniazid | Ethambutol | Streptomycin | Moxifloxacin | Ofloxaxin | Amikacin | Capreomycin | Kanamycin | Pyrazinamide | Ethionamide | Ciprofloxacin | Fluoroquinolones | PAS | Cycloserine | Delaminid |
|---|---|---|---|---|---|---|---|---|---|---|---|---|---|---|---|---|---|---|---|---|
| Sensitive | S1 | T | 2.2.1 | Beijing | S | S | S | S | S | S | S | S | S | S | S | S | S | S | S | S |
| | | M | 2.2.10 | n/a | S | S | S | S | S | S | S | S | S | S | NT | S | NT | NT | NT | NT |
| | S2 | T | 4.1.2.1 | T1 | S | S | S | rpsL p.Lys43Arg | S | S | S | S | S | S | S | S | S | S | S | S |
| | | M | 4.1.2.1 | n/a | S | S | S | rpsL p.Lys43Arg | S | S | S | S | S | S | NT | S | NT | NT | NT | NT |

| | | | | | | | | | | | | | | | | | | | | |
|---|---|---|---|---|---|---|---|---|---|---|---|---|---|---|---|---|---|---|---|---|
| | S3 | T | 4.1.1.1 | X2 | S | S | S | S | S | S | S | S | S | S | S | S | S | S | S | S |
| | S3 | M | 4.1.1.1 | n/a | S | S | S | S | S | S | S | S | S | S | NT | S | NT | NT | NT | NT |
| | S4 | T | 4.6 | Manu2 | S | S | S | S | S | S | S | S | S | S | S | S | S | S | S | S |
| | S4 | M | 3 and 4.6.2.2 | n/a | S | C in gene fabG1 | S | S | S | S | S | S | S | S | NT | S | NT | NT | NT | NT |
| | S5 | T | 4.3.4 | LAM9 | S | S | S | S | S | S | S | S | S | S | S | S | S | S | S | S |
| | S5 | M | 4.3.4.2 | n/a | S | S | S | S | S | S | S | S | S | S | NT | S | NT | NT | NT | NT |
| | S6 | T | 2.2.1 | Beijing | S | S | S | S | S | S | S | S | S | S | S | S | S | S | S | S |
| | S6 | M | 2.2 | n/a | S | S | S | S | S | S | S | S | S | S | NT | S | NT | NT | NT | NT |
| | S7 | T | 4.1.3 | T1 | S | S | S | S | S | S | S | S | S | S | S | S | S | S | S | S |
| | S7 | M | 4.1.3 | n/a | S | S | S | S | S | S | S | S | S | S | NT | S | NT | NT | NT | NT |
| | S8 | T | 1.2.1.2.1 | EAI2-nonthaburi | S | S | S | S | S | S | S | S | S | S | S | S | S | S | S | S |
| | S8 | M | 1.2.1 | n/a | S | S | S | S | S | S | S | S | S | S | S | S | S | S | S | S |
| Isoniazid mono resistance | I1 | T | 4.6.2.2 | Cameroon | S | fabG1 c.-15C>T | S | S | S | S | S | S | S | S | fabG1 c.-15C>T | S | S | S | S | S |
| | I1 | M | 4.6.2.2 | n/a | S | C in gene fabG1 | S | S | S | S | S | S | S | S | NT | S | NT | NT | NT | NT |
| | I2 | T | 4.6.1.2 | X1 | S | fabG1 c.-15C>T, inhA p.Ile194Thr | S | S | S | S | S | S | S | S | fabG1 c.-15C>T, inhA p.Ile194Thr | S | S | S | S | S |
| | I2 | M | 4.6.1.2 | n/a | S | inhA p.Ile194Thr C in gene fabG1 | S | S | S | S | S | S | S | S | NT | S | NT | NT | NT | NT |
| | I3 | T | 4.6.2.2 | Cameroon | S | fabG1 c.-15C>T | S | S | S | S | S | S | S | S | fabG1 c.-15C>T | S | S | S | S | S |
| | I3 | M | 4.6.2.2 | n/a | S | C in gene fabG1 | S | S | S | S | S | S | S | S | NT | S | NT | NT | NT | NT |
| | I4 | T | 4.6.2.2 | Cameroon | S | fabG1 c.-15C>T | S | S | S | S | S | S | S | S | fabG1 c.-15C>T | S | S | S | S | S |
| | I4 | M | 4.6.2.2 | n/a | S | C in gene fabG1 | S | S | S | S | S | S | S | S | NT | S | NT | NT | NT | NT |
| | I5 | T | 4.6.2.2 | Cameroon | S | fabG1 c.-15C>T | S | S | S | S | S | S | S | S | fabG1 c.-15C>T | S | S | S | S | S |

| | | | | | | | | | | | | | | | | | | | | |
|---|---|---|---|---|---|---|---|---|---|---|---|---|---|---|---|---|---|---|---|---|
| | | M | 4.6.2.2 | n/a | S | C in gene fabG1 | S | S | S | S | S | S | S | S | NT | S | NT | NT | NT | NT |
| MDR | M1 | T | 4.1.2.1 | H1 | rpoB p.Ser450Leu | katG p.Ser315Thr | M306I in gene embB | rpsL p.Lys43Arg | S | S | S | S | S | pncA p.Leu85Pro | S | S | S | S | S | S |
| | | M | 4.1.2.1 | n/a | rpoB p.Ser450Leu | katG p.Ser315Thr | M306I in gene embB | rpsL p.Lys43Arg | S | S | S | S | S | pncA p.Leu85Pro | NT | S | NT | NT | NT | NT |
| | M2 | T | 2.2.1 | Beijing | rpoB p.Ser450Leu, rpoB p.Glu761Asp | katG p.Ser315Thr | embB p.Asp354Ala | S | S | S | S | S | eis c.-37G>T | S | ethA c.-7T>C | S | S | S | S | S |
| | | M | 2.2.10 | n/a | rpoB p.Ser450Leu, rpoB p.Glu761Asp | katG p.Ser315Thr | embB p.Asp354Ala | S | S | S | S | S | S | S | NT | S | NT | NT | NT | NT |
| | M3 | T | 2.2.1 | Beijing | rpoB p.Ser450Leu, rpoB p.Glu761Asp | katG p.Ser315Thr | embB p.Asp354Ala | S | S | S | S | S | eis c.-37G>T | S | ethA c.-7T>C | S | S | S | S | S |
| | | M | 2.2.10 | n/a | S450L in gene rpoB | katG p.Ser315Thr | embB p.Asp354Ala | S | S | S | S | S | S | S | NT | S | NT | NT | NT | NT |
| | M4 | T | 4.2.1 | Ural-1 | rpoB p.Ser450Leu, rpoC p.Asp485Asn | inhA c.-154G>A, katG p.Ser315Thr | S | rpsL p.Lys88Arg | S | S | S | S | S | S | inhA c.-154G>A | S | S | S | S | S |
| | | M | 4.2.1 | n/a | S | katG p.Ser315Thr | S | S | S | S | S | S | S | S | NT | S | NT | NT | NT | NT |

T= TB-Profiler, M= Mykrobe, S= sensitive, NT = not tested

**S6 - Full ONT vs Illumina concordance table**

| | | | Lineage | Lineage name | Rifampicin | Isoniazid | Ethambutol | Streptomycin | Moxifloxacin | Ofloxacin | Amikacin | Capreomycin | Kanamycin | Pyrazinamide | Ethionamide | Ciprofloxacin | Fluoroquinolones | PAS | Cycloserine | Delaminid |
|---|---|---|---|---|---|---|---|---|---|---|---|---|---|---|---|---|---|---|---|---|
| Sensitive | S1 | P | n/a | n/a | S | S | S | S | NT | NT | NT | NT | NT | NT | NT | NT | NT | NT | NT | NT |
| | | O | 2.2.1 | Beijing | S | S | S | S | S | S | S | S | S | S | S | S | S | S | S | S |
| | | I | 2.2.1 | Beijing | S | S | S | S | S | S | S | S | S | S | S | S | S | S | S | S |
| | S2 | P | n/a | n/a | S | S | S | S | NT | NT | NT | NT | NT | NT | NT | NT | NT | NT | NT | NT |

| | | | | | | | | | | | | | | | | | | | | |
|---|---|---|---|---|---|---|---|---|---|---|---|---|---|---|---|---|---|---|---|---|
| | | O | 4.1.2.1 | T1 | S | S | S | rpsL p.Lys43Arg | S | S | S | S | S | S | S | S | S | S | S | S |
| | | I | 4.1.2.1 | T1 | S | S | S | rpsL p.Lys43Arg | S | S | S | S | S | S | S | S | S | S | S | S |
| | S3 | P | n/a | n/a | S | S | S | S | NT | NT | NT | NT | NT | NT | NT | NT | NT | NT | NT | NT |
| | | O | 4.1.1.1 | X2 | S | S | S | S | S | S | S | S | S | S | S | S | S | S | S | S |
| | | I | 4.1.1.1 | X2 | S | S | S | S | S | S | S | S | S | S | S | S | S | S | S | S |
| | S4 | P | n/a | n/a | S | S | S | S | NT | NT | NT | NT | NT | NT | NT | NT | NT | NT | NT | NT |
| | | O | 3 and 4.6 | CAS, Manu2 | S | S | S | S | S | S | S | S | S | S | S | S | S | S | S | S |
| | | I | 3 | CAS1-Delhi | S | S | S | S | S | S | S | S | S | S | S | S | S | S | S | S |
| | S5 | P | n/a | n/a | S | S | S | S | NT | NT | NT | NT | NT | NT | NT | NT | NT | NT | NT | NT |
| | | O | 4.3.4 | LAM9 | S | S | S | S | S | S | S | S | S | S | S | S | S | S | S | S |
| | | I | 4.3.4.2 | LAM9 | S | S | S | S | S | S | S | S | S | S | S | S | S | S | S | S |
| | S6 | P | n/a | n/a | S | S | S | S | NT | NT | NT | NT | NT | NT | NT | NT | NT | NT | NT | NT |
| | | O | 2.2.1 | Beijing | S | S | S | S | S | S | S | S | S | S | S | S | S | S | S | S |
| | | I | 2.2.1 | Beijing | S | S | S | S | S | S | S | S | S | S | S | S | S | S | S | S |
| | S7 | P | n/a | n/a | S | S | S | S | NT | NT | NT | NT | NT | NT | NT | NT | NT | NT | NT | NT |
| | | O | 4.1.3 | T1 | S | S | S | S | S | S | S | S | S | S | S | S | S | S | S | S |
| | | I | 4.1.3 | T1 | S | S | S | S | S | S | S | S | S | S | S | S | S | S | S | S |
| | S8 | P | n/a | n/a | S | S | S | S | NT | NT | NT | NT | NT | NT | NT | NT | NT | NT | NT | NT |
| | | O | 1.2.1.2.1 | EAI2-nonthaburi | S | S | S | S | S | S | S | S | S | S | S | S | S | S | S | S |
| | | I | 1.2.1.2.1 | EAI2-nonthaburi | S | S | S | S | S | S | S | S | S | S | S | S | S | S | S | S |
| Isoniazid mono | I1 | P | n/a | n/a | S | R | S | S | NT | NT | NT | NT | NT | NT | NT | NT | NT | NT | NT | NT |
| | | O | 4.6.2.2 | Cameroon | S | fabG1 c.-15C>T | S | S | S | S | S | S | S | S | fabG1 c.-15C>T | S | S | S | S | S |
| | | I | 4.6.2.2 | Cameroon | S | fabG1 c.-15C>T | S | S | S | S | S | S | S | S | fabG1 c.-15C>T | S | S | S | S | S |

| | | | | | | | | | | | | | | | | | | | | |
|---|---|---|---|---|---|---|---|---|---|---|---|---|---|---|---|---|---|---|---|---|
| | I2 | P | n/a | n/a | S | R | S | S | NT | NT | NT | NT | NT | NT | NT | NT | NT | NT | NT | NT |
| | I2 | O | 4.6.1.2 | X1 | S | fabG1 c.-15C>T, inhA p.Ile194Thr | S | gid c.102de1G | S | S | S | S | S | S | fabG1 c.-15C>T, inhA p.Ile194Thr | S | S | S | S | S |
| | I2 | I | 4.6.1.2 | X1 | S | fabG1 c.-15C>T, inhA p.Ile194Thr | S | gid c.102delG | S | S | S | S | S | S | fabG1 c.-15C>T, inhA p.Ile194Thr | S | S | S | S | S |
| | I3 | P | n/a | n/a | S | R | S | S | NT | NT | NT | NT | NT | NT | NT | NT | NT | NT | NT | NT |
| | I3 | O | 4.6.2.2 | Cameroon | S | fabG1 c.-15C>T | S | S | S | S | S | S | S | S | fabG1 c.-15C>T | S | S | S | S | S |
| | I3 | I | 4.6.2.2 | Cameroon | S | fabG1 c.-15C>T | S | S | S | S | S | S | S | S | fabG1 c.-15C>T | S | S | S | S | S |
| | I4 | P | n/a | n/a | S | R | S | S | NT | NT | NT | NT | NT | NT | NT | NT | NT | NT | NT | NT |
| | I4 | O | 4.6.2.2 | Cameroon | S | fabG1 c.-15C>T | S | S | S | S | S | S | S | S | fabG1 c.-15C>T | S | S | S | S | S |
| | I4 | I | 4.6.2.2 | Cameroon | S | fabG1 c.-15C>T | S | S | S | S | S | S | S | S | fabG1 c.-15C>T | S | S | S | S | S |
| | I5 | P | n/a | n/a | S | R | S | S | NT | NT | NT | NT | NT | NT | NT | NT | NT | NT | NT | NT |
| | I5 | O | 4.6.2.2 | Cameroon | S | fabG1 c.-15C>T | S | S | S | S | S | S | S | S | fabG1 c.-15C>T | S | S | S | S | S |
| | I5 | I | 4.6.2.2 | Cameroon | S | fabG1 c.-15C>T | S | S | S | S | S | S | S | S | fabG1 c.-15C>T | S | S | S | S | S |
| MDR | M1 | P | n/a | n/a | R | R | S | R | NT | NT | NT | NT | NT | R | NT | NT | NT | NT | NT | NT |
| MDR | M1 | O | 4.1.2.1 | H1 | rpoB p.Ser450Leu | katG p.Ser315Thr | embB p.Met306Ile | rpsL p.Lys43Arg | S | S | S | S | S | pncA p.Leu85Pro | S | S | S | S | S | S |
| MDR | M1 | I | 4.1.2.1 | H1 | rpoB p.Ser450Leu | katG p.Ser315Thr | embB p.Met306Ile | rpsL p.Lys43Arg | S | S | S | S | S | pncA p.Leu85Pro | S | S | S | S | S | S |
| MDR | M2 | P | n/a | n/a | R | R | S | R | NT | NT | NT | NT | NT | R | NT | NT | NT | NT | NT | NT |
| MDR | M2 | O | 2.2.1 | Beijing | rpoB p.Ser450Leu, rpoB p.Glu761Asp | katG p.Ser315Thr | embB p.Asp354Ala | S | S | S | S | S | eis c.-37G>T | S | ethA c.-7T>C | S | S | S | S | S |
| MDR | M2 | I | 2.2.1 | Beijing | rpoB p.Ser450Leu, rpoB p.Glu761Asp | katG p.Ser315Thr | embB p.Asp354Ala | S | S | S | S | S | eis c.-37G>T | S | ethA c.-7T>C | S | S | S | S | S |
| MDR | M3 | P | n/a | n/a | R | R | S | S | NT | NT | NT | NT | NT | R | NT | NT | NT | NT | NT | NT |
| MDR | M3 | O | 2.2.1 | Beijing | rpoB p.Ser450Leu, rpoB p.Glu761Asp | katG p.Ser315Thr | embB p.Asp354Ala | S | S | S | S | S | eis c.-37G>T | S | ethA c.-7T>C | S | S | S | S | S |

| | | | | | | | | | | | | | | | | | | | |
|---|---|---|---|---|---|---|---|---|---|---|---|---|---|---|---|---|---|---|---|
| | I | 2.2.1 | Beijing | rpoB p.Ser450Leu, rpoB p.Glu761Asp | katG p.Ser315Thr | embB p.Asp354Ala | S | S | S | S | S | eis c.-37G>T | S | ethA c.-7T>C | S | S | S | S | S |
| M4 | P | n/a | n/a | R | R | S | R | NT | NT | NT | NT | NT | S | NT | NT | NT | NT | NT | NT |
| | O | 4.2.1 | Ural-1 | rpoB p.Ser450Leu, rpoC p.Asp485Asn | inhA c.-154G>A, katG p.Ser315Thr | S | rpsL p.Lys88Arg | S | S | S | S | S | S | inhA c.-154G>A | S | S | S | S | S |
| | I | 4.2.1 | Ural-1 | rpoB p.Ser450Leu, rpoC p.Asp485Asn | inhA c.-154G>A, katG p.Ser315Thr | S | rpsL p.Lys88Arg | S | S | S | S | S | S | inhA c.-154G>A | S | S | S | S | S |

P = phenotypic, O = Oxford Nanopore Technologies WGS, I = Illumina WGS, NT = Not tested, R = resistant, S = sensitive. Only resistance mutations of final confidence grading of ≥3 according to the WHO's Catalogue of mutations in Mycobacterium tuberculosis complex and their association with drug resistance.
